# Identification of Potential Therapeutic targets for Stroke and Its Subtypes by Integrating Proteomes and Genetics from Human Plasma

**DOI:** 10.1101/2023.08.29.23294808

**Authors:** Hanchen Liu, Xiaoxi Zhang, Hongyu Ma, Thanh N. Nguyen, Weilong Hua, Shaojun Mo, Qinghai Huang, Jianmin Liu, Yu Zhou, Pengfei Yang

**Affiliations:** Neurovascular Center, Naval Medical University Changhai Hospital, Shanghai, China; Boston University Chobanian and Avedisian School of Medicine, Department of Neurology & Radiology, Boston Med Center, Boston, MA 02118 USA

**Keywords:** stroke, Mendelian randomization, Proteome-wide association study, Transcriptome-wide association study, Bayesian colocalization, Human plasma proteomes

## Abstract

**Background:** Previous genome-wide association studies (GWAS) have identified several risk genes for stroke; however, it remains unclear how they confer risk for the disease. We conducted an integrative analysis to identify candidate genes for stroke and stroke subtypes by integrating blood-derived multi-omics data with genetic data.

**Method:** We systematically integrated the latest stroke GWAS database (73,652 patients and 1,234,808 controls) with human plasma proteomes (N=7,213) and performed proteome-wide association studies (PWAS), Mendelian randomization (MR), Bayesian colocalization analysis, and transcriptome-wide association study (TWAS) to prioritize genes that associate the risk of stroke and its subtypes with their expression and protein abundance in plasma. Cell-type specificity and functional enrichment analysis using single-cell RNA sequencing (scRNA-seq) and Gene Ontology (GO) databases were then performed to select target genes. A two-step MR analysis was followed to explore the potential mechanisms.

**Results:** We found that the protein abundance of seven genes (*MMP12, F11, SH3BGRL3, ENGASE, SCARA5, SWAP70,* and *SPATA20*) in the plasma was associated with stroke and its subtypes, with six genes (*MMP12, F11, SH3BGRL3, SCARA5, SWAP70,* and *SPATA20*) causally related with stroke and its subtypes (P < 0.05/proteins identified for PWAS; P < 0.05/8 for MR; posterior probability of hypothesis 4 ≥ 75 % for Bayesian colocalization). The effect of *F11, SH3BGRL, SPATA20,* and *SWAP70* on each subtype was mediated by Factor XI inhibitors (FXI), atrial fibrillation, T2D, and SBP respectively (p<0.05). We also found that *SCARA5* and *SWAP70* were related to stroke and ischemic stroke at the transcriptome level.

**Conclusions:** Our present proteomic findings have identified new causal genes in the pathogenesis of stroke, which may offer potential future therapeutic targets for stroke prevention.

## INTRODUCTION

Stroke is the leading cause of death and long-term disability worldwide, which imposes a great burden on global health.^1^ Despite significant advancements in the past decades, critical gaps persist in the management of stroke and its subtypes(i.e. large artery atherosclerosis, cardioembolic stroke, or small artery occlusion).^2^ As such, better understanding the mechanism of stroke occurrence, and identifying novel therapeutic targets are needed to tackle this challenge.

In recent years, large-scale genome-wide association studies (GWAS) have greatly aided the discovery of genetic loci linked to stroke and its subtypes.^3^ However, despite this progress in understanding the genetic risk of stroke, genes undergo regulation at the post-transcriptional, translational, and post-translational levels, predisposition to stroke involves a complex, pleiotropic, and polygenic genetic architecture. Deciphering the underlying mechanism for these genetic effects on stroke is always challenging, which hinders the translation of identified genetic findings into drug development.

Proteins are the final products of gene expression and the main functional components of cells and biological processes.^4,5^ Directly investigating proteins that are linked to stroke would be of great help in identifying potential therapeutic targets regarding stroke management, especially when this link is supported by the genetic association.^6^ In the past few years, several genetic articles concerned the role of circulating proteins in stroke, including a proteome-wide association study and one Mendelian randomization (MR) analysis.^7,8^ However, it is likely that the identification of protein quantitative trait locus (pQTL) in their study was incomplete, both due to inadequate sample size or/and lack of comprehensive protein measurements, and also multi-omics verification was lacking. With higher-quality and larger sample sizes of GWAS and, especially, human plasma proteomics data, becoming available, it would be promising to perform a new analysis using the newest data and tools to identify novel therapeutic targets for stroke.

Accordingly, we conducted an integrative analysis to identify candidate genes for stroke and stroke subtypes by integrating blood-derived multi-omics data with genetic data.

## METHODS

### Human plasma proteomic and transcriptomic data

The plasma proteomic data were derived from a comprehensive set of analyses of *cis*-genetic regulation of the plasma proteome in large European cohorts of the Atherosclerosis Risk in Communities (ARIC) study, comprising cleaned plasma protein data of 7,213 European American participants.^9^ Relative concentrations of plasma proteins or protein complexes were measured by the Slow-Of rate Modified Aptamer (SOMAmer) via a proteomic profiling platform.^10,11^ 4657 SOMAmers were analyzed, which tagged proteins or protein complexes encoded by 4,435 genes. Finally, a total of 2,004 significant SOMAmers were identified.

The whole-blood eQTL data was derived from the Young Finns Study (YFS), in which the expression of peripheral blood genes (n=1264) was comprehensively analyzed.^12^

### GWAS data of stroke and its subtypes

Genetic association estimates for stroke were obtained from the GIGASTROKE consortium, the largest published stroke GWAS meta-analysis to date.^13^ Populations were restricted to European participants. Data for 1,308,064 European descent individuals (73,652 any stroke cases and 1,234,808 controls) were used as a stroke GWAS summary dataset. The primary outcomes for main analysis were the occurrence of any stroke (including both ischemic and hemorrhagic stroke; AS; 73,652 cases), any ischemic stroke (AIS; 62,100 cases), or three aetiologic ischemic stroke subtypes: large artery stroke (LAS; 6,399 cases), cardioembolic stroke (CES; 10,804 cases) and small vessel stroke (SVS; 6,811 cases).

### GWAS data on risk factors for stroke

The secondary outcomes we considered were stroke risk factors, which were selected from a literature review.^14^ Five risk factors were considered for the two-sample MR analyses and we sought well-powered publicly available GWAS summary statistics for these risk factors, including systolic blood pressure (SBP),^15^ atrial fibrillation (AF),^16^ type 2 diabetes (T2D),^17^ body mass index (BMI),^18^ and coagulation factor XI (FXI)^19^. The full details of the data sources and sample size for these GWASs were provided in supplementary materials (**Table S1**).

#### The RNA-Sequencing Data Availability of stroke

Single-cell RNA sequencing (scRNA-seq) data were used to identify the cell-type specificity and functional pathways of target genes.^20^ The scRNA-seq data were downloaded from the GEO dataset (https://www.ncbi.nlm.nih.gov/geo/) with accession number GSE174574. This study provided a scRNA-seq landscape of 17 cell populations of mice brain cortex with cell-type specific gene expression profiles based on the middle cerebral artery occlusion (MCAO) model.^20^ Functional pathways of target genes were enriched by Gene ontology (GO) analysis. Cell type-specific expression of the risk genes was displayed with a dot plot according to the cell classification from Zhang et al.^20^ The scRNA-seq analysis was performed using R packages of Seurat and ggsci. The enrichment score of risk genes from PWAS was calculated by R package AUCell.

### Statistical analysis

#### Proteome-wide and transcriptome-wide association studies

The overall design of this study is shown in **Figure 1**. Proteome-wide association studies (PWAS) and transcriptome-wide association studies (TWAS) were carried out using FUSION.^21^ A linkage disequilibrium (LD) reference panel was used to minimize the influence of LD on the estimated test statistics.^21^ We used FUSION to compute the effect of single nucleotide polymorphisms (SNPs) on protein abundance for proteins with significant heritability (heritability P < 0.01). Two predictive models, enet and top1 were adopted in the analysis.^9^ Protein weights from the most predictive model were selected,^9^ and the expression weights were derived from transcriptomic data generated from YFS. Subsequently, we used FUSION to combine the genetic effect of stroke and stroke subtypes (stroke and its subtypes GWAS z-score) with the protein or expression weights by calculating the linear sum of z-score × weight for the independent SNPs at the locus to perform the PWAS or TWAS. The P values were adjusted for false discovery rate (FDR) using Benjamini-Hochberg (BH) method and the statistical significance was set at P<0.05.

**Figure 1.**
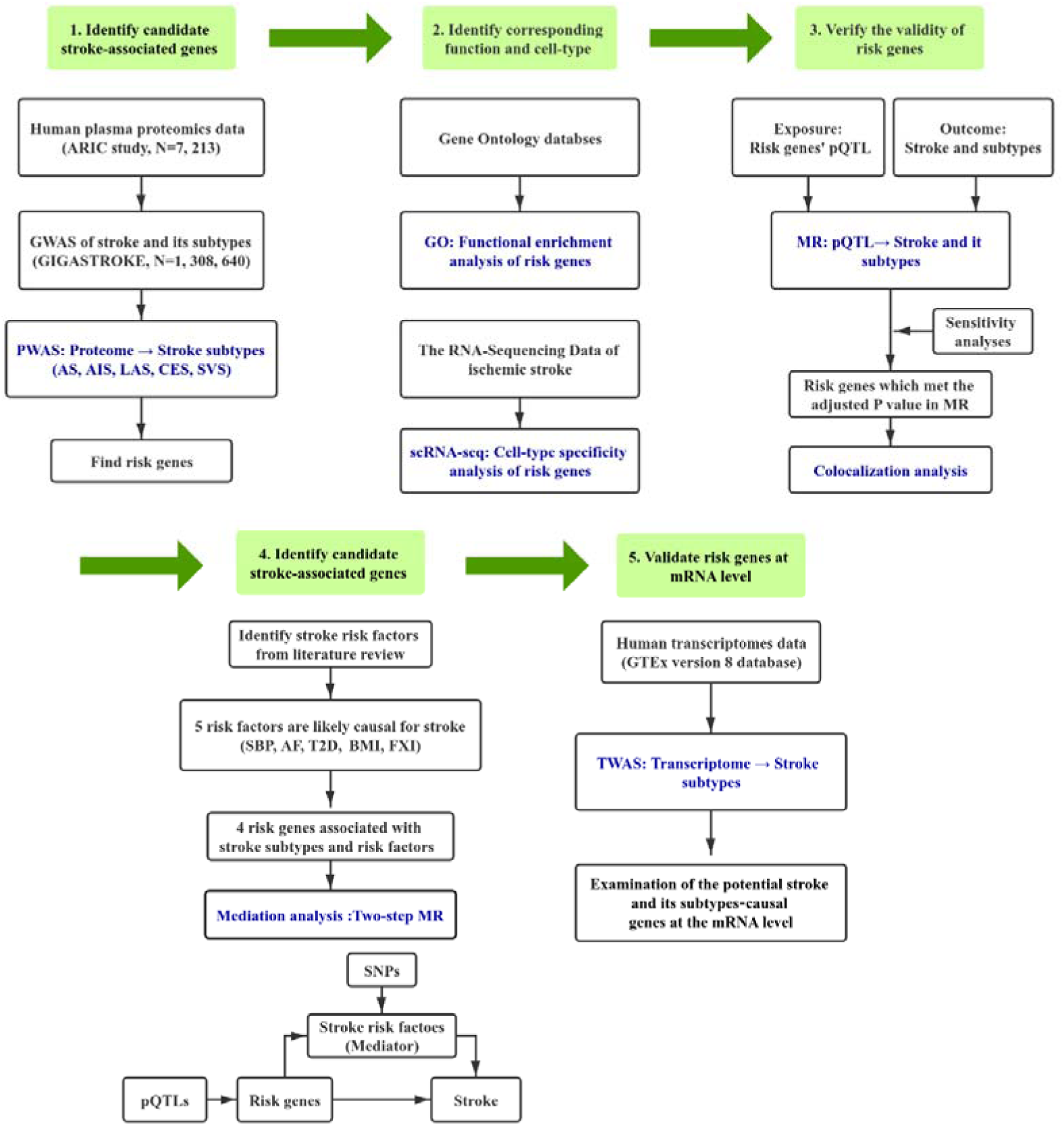
Overview of this study. First, we used protein quantitative trait locus (pQTL) datasets obtained from plasma and findings from stroke and its subtypes GWAS to perform a PWAS analysis. Second, we performed cell enrichment and functional enrichment analysis of the identified risk genes using single-cell transcriptomes and Gene Ontology (GO) databases. Third, we used independent Mendelian randomization (MR) analysis to verify PWAS-significant genes. Fourth, we used a colocalization to integrate GWAS data and plasma pQTL using a Bayesian colocalization analysis to explore whether two associated signals are consistent with shared causal variant(s). Fifth, we included five common risk factors for stroke in two-step MR analyses to explore the clinical mechanisms of risk genes. Finally, we explored the significant genes driving GWAS signals at the transcriptional level by leveraging gene expression data. ARIC, Atherosclerosis Risk in Communities; GWAS, genome-wide association studies; PWAS, proteome-wide association studies; AS: any stroke; AIS: any ischemic stroke; LAS: large artery stroke; CES: cardioembolic stroke; SVS: small vessel stroke; scRNA-seq, single-cell RNA sequencing; pQTL: protein quantitative trait locus; MR, Mendelian randomization; SBP, systolic blood pressure; AF, atrial fibrillation; T2D, type 2 diabetes; BMI, body mass index; FXI, coagulation factor XI; GTEx, Genotype-Tissue Expression; TWAS, transcriptome-wide association study.

#### Mendelian Randomization analysis

We used two-sample MR to verify whether stroke and its subtypes PWAS-significant genes (from the FUSION approach) were associated with stroke and its subtypes via their cis-regulated plasma protein abundance and estimate the role of protein expressed by these genes in stroke subtypes and its risk factors. The SNPs included in the study robustly and independently (r^2^ < 0.1; clumping window, 10 000 kb) predicted exposures at a genome-wide level (P < 5 × 10^-8^). We harmonized the SNP alleles across studies and removed palindromic SNPs with ambiguous allele frequencies (0.42–0.58).

Principal analyses were conducted using the inverse-variance weighted (IVW) MR method. This method assumes that all genetic variants are valid instrumental variables and produces the most accurate estimates.^22^ Wald’s ratio method was applied when there was only one SNP available for the target exposure.^23^ The weighted median and MR-Egger regression were adopted as sensitivity analyses. The weighted median approaches give more weight to the instrumental variables that are more precise, and the estimate is consistent even when up to 50 % of the information comes from invalid or weak instruments.^24^ The MR-Egger could detect and adjust for directional pleiotropy, albeit with low precision.^22^ We used Cochran’s Q statistic in the IVW model to assess the heterogeneity between variant-specific estimates. The MR Pleiotropy Residual Sum and Outlier (PRESSO) approaches were used to identify possible outliers. Finally, a leave-one-SNP-out analysis was performed in which SNPs were systematically removed to assess whether a single SNP drove the results. We set the P value for statistical significance at P<0.007, after correcting for multiple exposures using the Bonferroni method (α=0.05/7 proteins). Statistical analyses were conducted in R (version 4.2.2) and MR analyses were conducted using “TwoSampleMR”.

#### Bayesian colocalization analysis

We conducted colocalization analysis for each of the genes associated with one or more of the stroke outcomes to investigate whether the protein levels of a gene and stroke outcome genetic associations are due to the same causal variants. We estimated the posterior probability (PP) of multiple traits sharing the same causal SNP simultaneously using a multi-trait colocalization method.^25^ We used the default colocalization priors of P_1_ = 10^−4^, P_2_ = 10^−4^, and P_12_ = 10^−5^, where P_1_ is the probability that a given variant is associated with stroke or its subtypes, P_2_ is the probability that a given variant is a significant pQTL, and P_12_ is the probability that a given variant is both a stroke or its subtypes result and a pQTL. colocalization uses computed approximation Bayes factors and summary association data to generate a posterior probability for the following 5 hypotheses: H_0_, No association with either GWAS or pQTL; H_1_, Association with GWAS, not with pQTL; H_2_, Association with pQTL, not with GWAS; H_3_, Association with GWAS and pQTL, two independent SNPs; and H_4_, Association with GWAS and pQTL, one shared SNP. We mainly focused on the last hypothesis H4 and posterior probability (PP) was used to quantify support for H4 (denoted as PPH4). We defined strong evidence of colocalization at PPH4 ≥ 0.75.^26^

#### Mediation analysis

For protein levels of a gene that causally associate with both stroke and risk factors, we conducted a mediation analysis to quantify the effects of protein levels of a gene on stroke outcomes via risk factors. The “total” effect of exposure on outcome includes both the “direct” effect and any “indirect” effect via one or more mediators. In this study, the total effect is captured by a standard univariable MR analysis—the primary MR. To decompose direct and indirect effects, we used results from two-step MR and chose the Product method to estimate the beta of indirect effect and the Delta method to estimate the standard error (SE) and confidence interval (CI).^27^

#### Functional enrichment analysis and Cell-type specificity analysis

Gene ontology (GO) analysis was performed to do functional enrichment analysis. Subsequently, using mouse single-cell RNA-seq data of the CoW profiled, we investigated the cell type-specific expression of the risk genes. The cell suspensions underwent scRNAseq using standard 10X Chromium Single Cell Chemistry V3. Raw sequencing data were processed using Cellranger to produce gene-level counts for each cell in each sample. The CellRanger mkfastq command was used to generate Fastq files. Data were mapped to a prebuild mouse reference genome. All subsequent analysis was performed using R packages of Seurat and ggsci. The enrichment score of risk genes from PWAS was calculated by R package AUCell. The detailed parameters of scRNA analysis could refer to previous works.^20^

## RESULTS

### PWAS of stroke and its subtypes

The PWAS identified 7 genes (*MMP12*, *F11*, *ENGASE*, *SH3BGRL3*, *SPATA20*, *SWAP70*, *SCARA5*) whose cis-regulated plasma protein levels were associated with stroke and its subtypes at a false discovery rate (FDR) of P<0.05 (**Figure 2 and Table S2**). The protein abundances of *MMP12* were associated with AS, AIS, and LAS (AS: Z-score: -5.881, PWAS FDR P = 2.73×10^-6^; AIS: Z-score: -6.149, PWAS FDP P= 5.18×10^-7^; LAS: Z-score: -5.674, PWAS FDR P= 7.41×10^-6^), and the protein abundances of *F11* was associated with AS, AIS, and CES (AS: Z-score: 5.972, PWAS FDR P = 3.12×10^-6^; AIS: Z-score: 6.718, PWAS FDR P = 2.45×10^-8^; CES: Z-score: 5.683, PWAS FDR P = 1.39×10^-5^). The protein abundances of *ENGASE* and *SH3BGRL3* were associated with AS and AIS (*ENGASE*: AS: Z-score= 5.055, PWAS FDR P = 1.91×10^-4^; AIS: Z-score= 4.805, PWAS FDR P = 6.87×10^-4^), (*SH3BGRL3*: AS: Z-score= -4.284, PWAS FDR P = 6.14×10^-3^; AIS: Z-score= -4.322, PWAS FDR P = 5.12×10^-3^). The protein abundances of *SCARA5*, *SWAP70* and *SPATA20* were associated with AS, AIS and SVS respectively (*SCARA5*: Z-score: -3.712, PWAS FDR P = 4.58×10^-2^; *SWAP70*: Z-score: -3.955, PWAS FDR P = 2.03×10^-2^; *SPATA20*: Z-score: 4.55, PWAS FDR P = 5.21×10^-3^). (**Table S1**)

**Figure 2.**
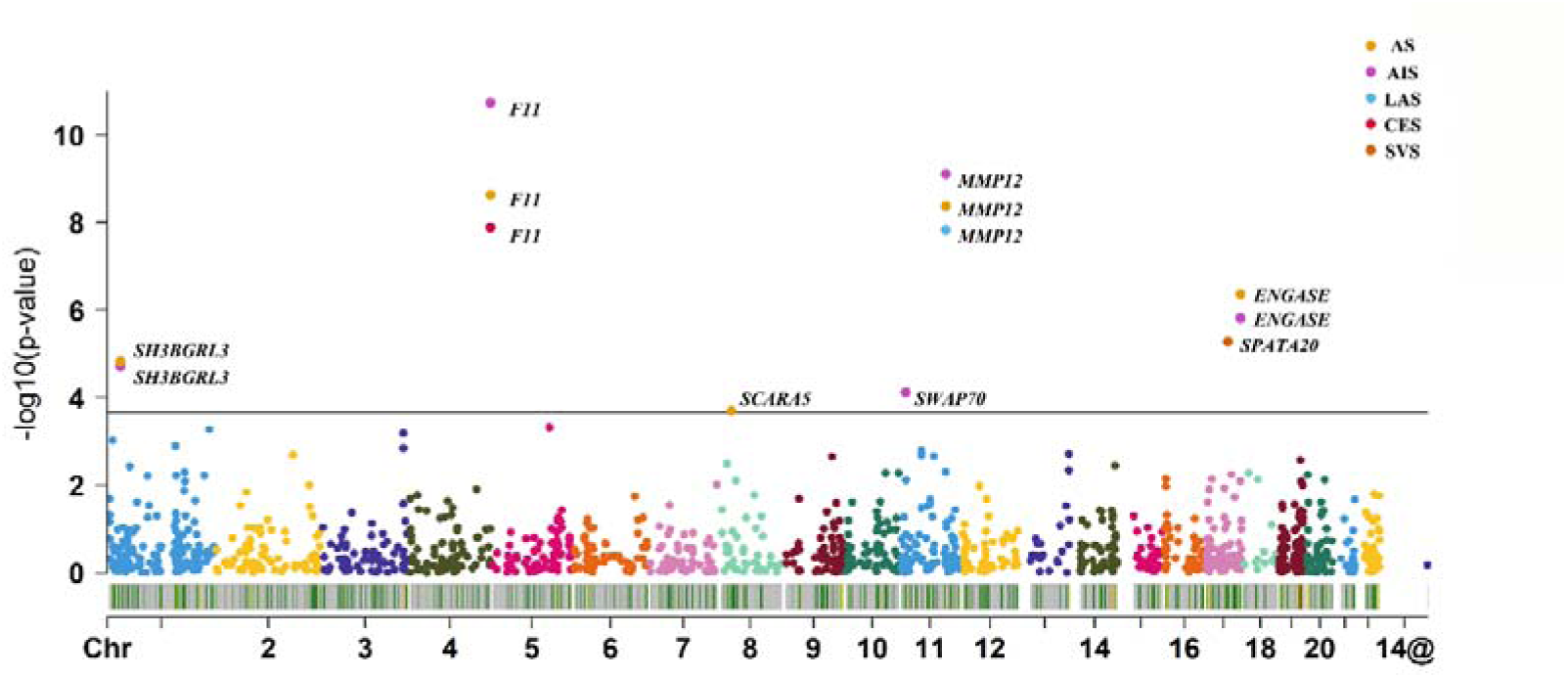
The Manhattan plot for the PWAS of stroke and stroke subtypes. The Manhattan plot shows the genes identified using PWAS for AS, AIS, LAS, CES, and SVS, respectively. Each point represents a single association test between a gene and stroke and its subtypes ordered by genomic position on the x axis and the association strength on the y axis as the −log10(P) of a z-score test. The discovery PWAS identified 7 genes whose cis-regulated plasma protein abundance was associated with stroke subtypes at an FDR of P < 0.05. The red horizontal line reflects the significant threshold of the FDR P < 0.05.

### Functional enrichment analysis and Cell-type specificity analysis

We investigated whether the risk genes identified by PWAS were enriched in some particular functions and some particular brain cell types. Go analyses suggest that *MMP12* and *F11* were significantly related to serine-type endopeptidase activity, as well as serine-type peptidase activity, wound healing, and serine hydrolase activity et.al. *MMP12* and *SWAP70* were significantly related to negative regulation of cell adhesion and positive regulation of response to external stimulus. *MMP12* and *ENGASE* were significantly related to the glycoprotein metabolic process. *SWAP70* and *SH3BGRL3* were significantly related to the cell leading edge (**Figure 3A**). Single-cell RNA-seq data showed different expression patterns of the risk genes in the brain. *MMP12* was mainly expressed in microglia. *F11* was mainly expressed in fibroblast. *SCARA5* was mainly expressed in perivascular fibroblast-like cells and *SWAP70* was mainly expressed in endothelial. *SH3BGRL3* was abundantly expressed in most cell types of the brain. The expression of *SPATA20* was not detected in this system (**Figure 3B** and **Table S3**).

**Figure 3.**
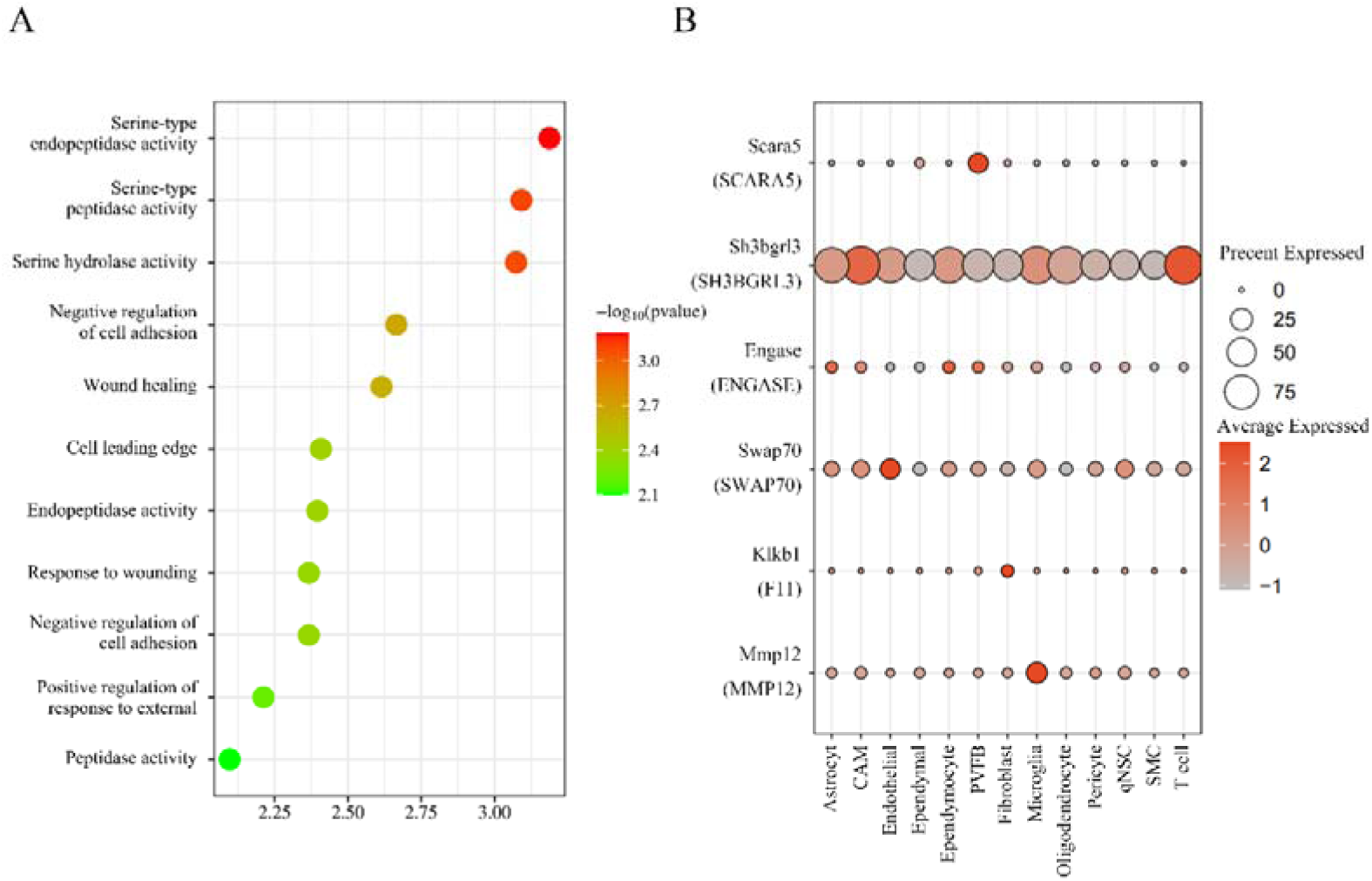
The functional enrichment analysis and Cell-type specificity analysis among candidate genes. A: functional enrichment analysis; B: cell-type specificity analysis, Single-cell-type expression of the potentially stroke-risk genes. Bar graph of single-cell-type enrichment for risk genes in stroke and its subtypes from the discovery PWAS. The diagram depicts expression of every gene (y axis) for each cell (x axis).

### Genetically determined PWAS-significant genes and risk of stroke and its subtypes

Cis-regulated plasma protein levels of seven PWAS-significant genes were tested for causal relationships with stroke and its subtypes by using MR analysis (P<0.007 = 0.05/7). As shown in **Figure 4**, we found that the concentration of *MMP12* was inversely associated with AS, AIS, and LAS risk (AS: OR [95% CI]: 0.93 [0.91, 0.95]; AIS: OR [95% CI]: 0.92 [0.91, 0.94]; LAS: OR [95% CI]: 0.82 [0.75, 0.91]), while *F11* was positively associated with AS, AIS, and CES risk (AS: OR [95% CI]: 1.07 [1.05, 1.10]; AIS: OR [95% CI]: 1.09 [1.06, 1.13]; CES: OR [95% CI]: 1.23 [1.15, 1.31]). The concentration of *ENGASE* was positively associated with AS and AIS (AS: OR [95% CI]: 1.05 [1.03, 1.06]; AIS: OR [95% CI]: 1.04 [1.03, 1.06]), but the concentration of *SH3BGRL3* was inversely associated with AS, AIS (AS: OR [95% CI]: 0.96 [0.94, 0.98]; AIS: OR [95% CI]: 0.95 [0.93, 0.97]). *SCARA5* and *SWAP70* were associated with a lower risk of AS (OR [95% CI]: 0.92 [0.87, 0.98]) and AIS (OR [95% CI]: 0.96 [0.94, 0.99]) while *SPATA20* was associated with a higher risk of SVS (OR [95% CI]: 1.16 [1.09, 1.23]). All sensitivity analyses supported the main results of the MR Analysis (**Table S4**). The results of MR further confirmed the association between seven genes and their corresponding stroke subtypes.

**Figure 4.**
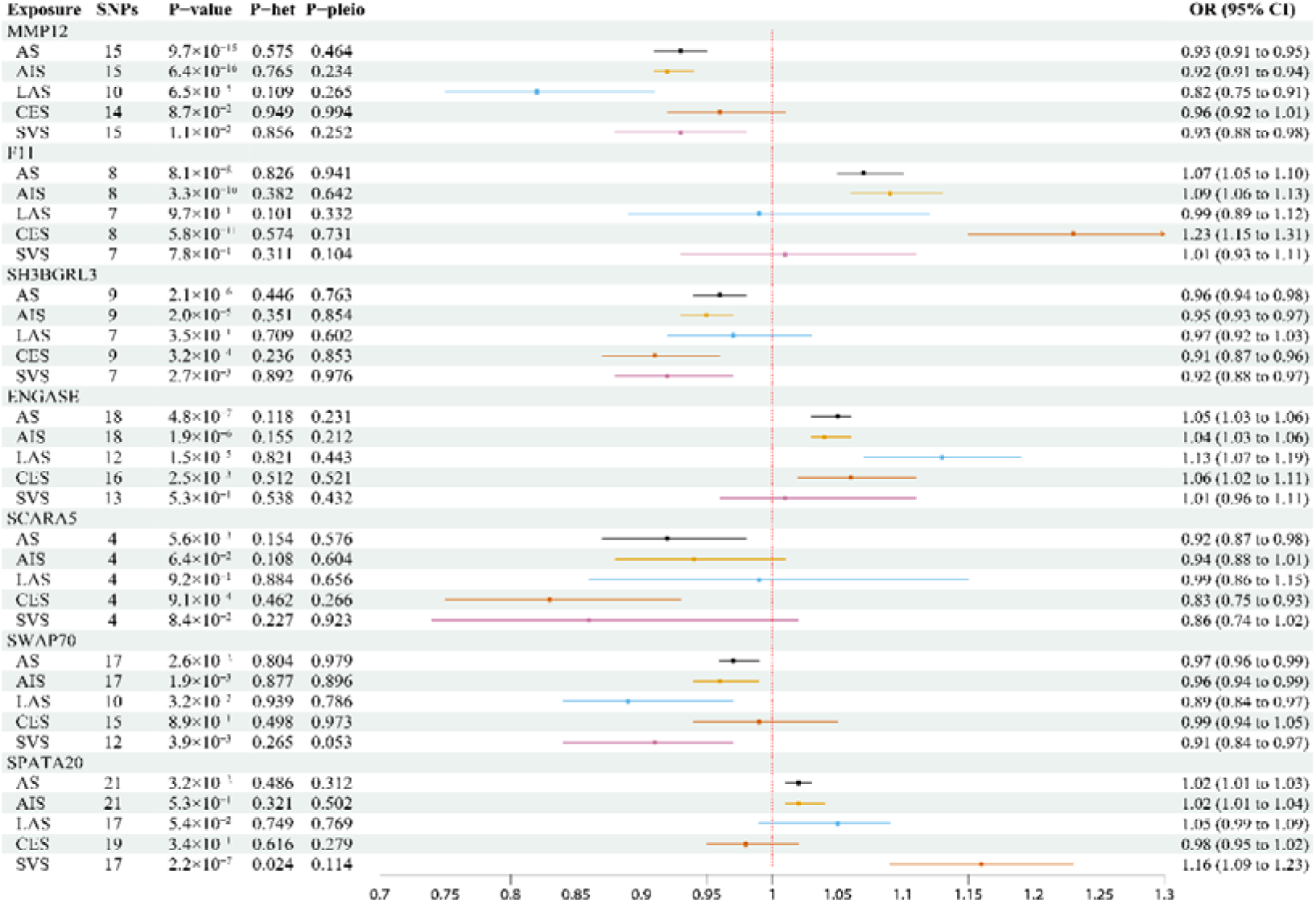
Effects of stroke and its subtypes PWAS-significant genes on stroke subtypes. The squares are the causal estimates on the OR scale, and the whiskers represent the 95% confidence intervals for these ORs. SNPs: number of SNPs used for the estimation of the causal effects in this plot. P-value were determined from the inverse-variance-weighted two-sample MR method. P-het, P value in the Q statistic for heterogeneity; P-pleio, P value in the Egger intercept.

### Colocalization between stroke risk genes and pQTLs in the plasma

We examined the PP for a shared causal variant between a pQTL and stroke for the seven genes which met the corrected P value threshold in previous PWAS and MR analysis (**Table S10**). The colocalization suggested the genetic variants associated with *MMP12* (pQTL) were due to the same genetic variants underlying the association with AS, AIS, and LAS (PPH4 ≥ 0.75). Similarly, *F11* pQTLs colocalized with AS, AIS, and CES genetic associations; *SH3BGRL3* pQTLs colocalized with AS and AIS; *SCARA5*, *SWAP70*, and *SPATA20* pQTLs colocalized with AS, AIS, and SVS respectively. It suggests that six of seven proteins play an important role in the pathophysiology of stroke subtypes.

### Significance of the protein findings

To determine the importance of the six of seven (*ENGASE* was excluded because its results of Bayesian colocalization analysis were negative) potentially causal genes identified from the PWAS, MR, and Bayesian colocalization analyses, we obtained the lowest P values for the SNPs within 1 Mb of each of these seven genes using the summary statistics from the most extensive stroke GWAS (N=446, 696).^28^ The most significant P values were less than 5×10^-8^ in two genes (*MMP12* and *F11*), while the P values of SNPs in the remaining 4 genes ranged from 1.32×10^-5^ to 2.36×10^-6^ (**Table S5**). The findings suggest that specific plasma proteins likely contribute to the pathogenesis of stroke subtypes.

### Mediation Effect of six genes on stroke subtypes via risk factors

To investigate the indirect effect of cis-regulated plasma protein levels of six genes on stroke subtypes via risk factors, we carried out a mediation analysis using the effect estimates from two-step MR and the total effect from primary MR. We calculated the causal relationship between five risk factors and stroke using MR Analysis (**Table S6**).

This analysis was restricted to four genes, *F11*, *SWAP70*, *SH3BGRL3*, and *SPATA20*, that showed evidence of an effect in both MRs with risk factors and stroke outcomes (**Table S6-S7**). The mediation effect of *F11* via FXI is the highest (63.8%). The indirect effect of *SPATA20* on the risk of SVS via T2D contributes to one-fourth of the total effect (25.2%). Similarly, the proportion of the mediation effect of *SWAP70* and *SH3BGRL3* on AIS and AS via SBP and AF is one-tenth, respectively (*SWAP70*: 14.1%; *SH3BGRL3*: 12.2%). (**Figure 5**).

**Figure 5.**
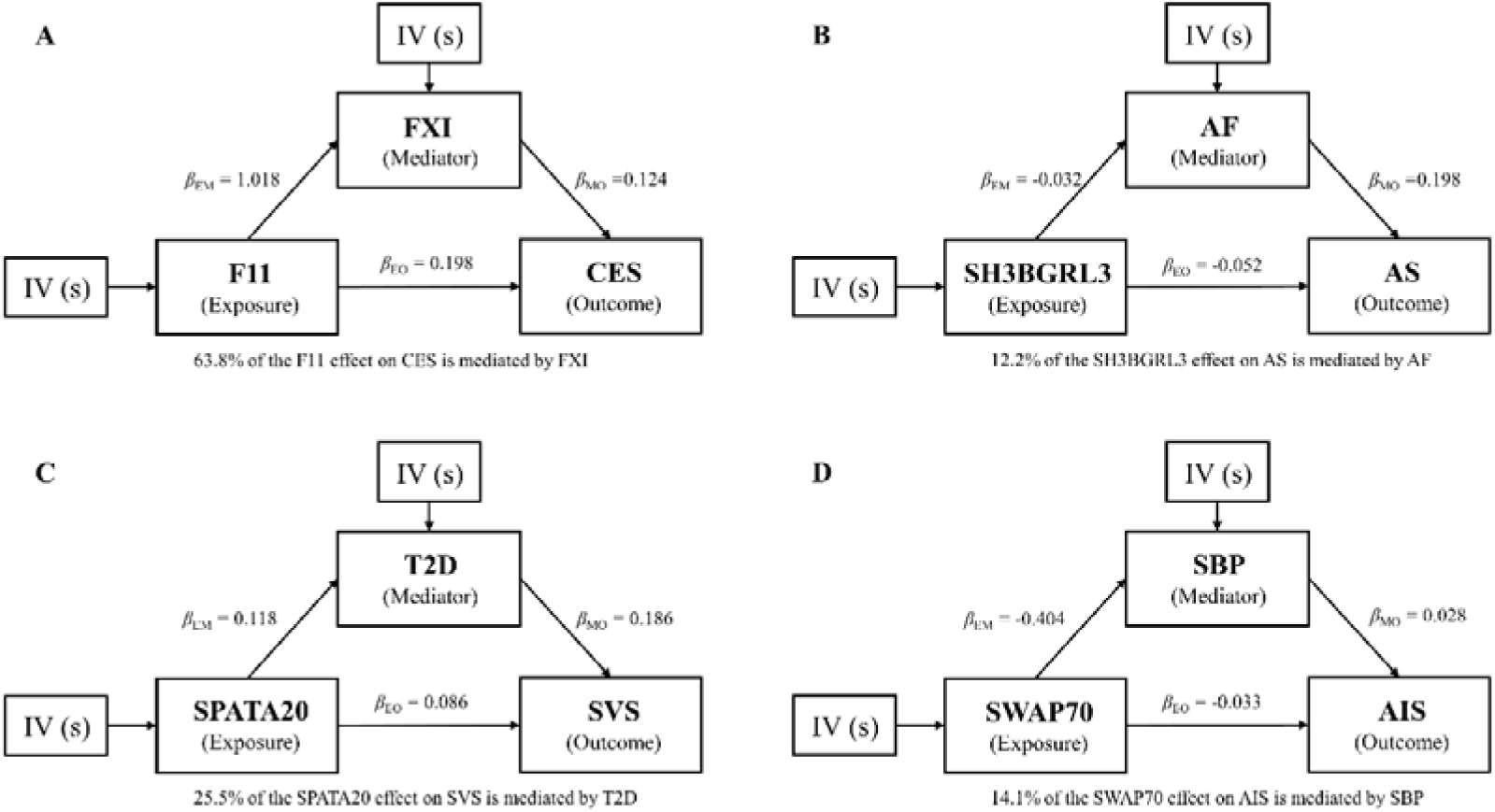
Mediation effects of genes on stroke via risk factors. Mediation analyses to quantify the effects of four genes on stroke subtypes via risk factors. **A**. *F11* effect on CES mediated by FXI. **B.** *SH3BGRL3* effect on AS mediated by AF. **C**. *SPATA20* effect on SVS mediated by T2D. **D**. *SWAP70* effect on AIS mediated by SBP. FXI, coagulation factor XI; CES, cardioembolic stroke; SBP, systolic blood pressure; AF, atrial fibrillation; T2D, type 2 diabetes; SVS, small vessel stroke; AS, any stroke; AIS, any ischemic stroke.

### Examination of the potential stroke-causal proteins at the mRNA level

We combined the stroke subtypes GWAS data with human plasma transcriptomes to conduct stroke and its subtypes TWAS using FUSION. We found that the cis-regulated plasma mRNA expression of the forty-one genes was associated with stroke and its subtypes (FDR P < 0.05) (**Table S8**). Interestingly, we found two genes identified in TWAS, i.e. *SWAP70* and *SPATA20*, were also significant in the AIS and SVS PWAS respectively, suggesting joint evidence from PWAS and TWAS for its role in stroke etiology.

## DISCUSSION

In the present study, we employed an integrated analysis based on GWAS, PWAS, TWAS, MR, and Bayesian colocalization to investigate the causal relationship between proteins in the plasma and stroke and its subtypes. We identified 7 potential risk genes (*MMP12, F11, SH3BGRL3, ENGASE, SCARA5, SWAP70,* and *SPATA20*) of stroke with altered protein abundances in the plasma. We enriched all seven genes into corresponding functional pathways and cell types. Six (*MMP12, F11, SH3BGRL3, SCARA5, SWAP70,* and *SPATA20*) of the above genes were replicated in the independent MR and Bayesian colocalization analysis validation analyses of stroke and its subtypes, providing a higher confidence level. Furthermore, *SWAP70* and *SPATA20* were supported at the transcriptional level. Also, through a mediation analysis, we found the effect of *F11* on CES, *SH3BGRL3* on AS and AIS, *SPATA20* on SVS, and *SWAP70* on AIS were partly mediated by FXI, AF, T2D, and SBP respectively, demonstrating a critical role of these risk factors in the pathogenesis of stroke in consistence with previous epidemiological studies.^29–33^

Of the 7 genes studied, three (*MMP12*, *F11*, and *SCARA5*) have been implicated in previous articles.^7,8^ *MMP12* is a member of the matrix metalloproteinase family that contributes to vascular remodeling. To date, the relationship between its circulating level and stroke is unclear and contradicted in previous studies. In one population-based study containing 2983 participants, higher *MMP12* level was independently associated with ischemic stroke risk (HR=1.30, 95% CI 1.16-1.45, P=4.55×10^-06^).^34^ In contrast, our study is consistent with another genetic study which showed an inverse relationship between *MMP12* circulating level and risk of ischemic stroke.^35^ FXI is mediated by the *F11* gene and has become increasingly interesting for its role in the pathogenesis of thrombosis. A GWAS analysis containing 371,695 participants found a lower FXI level was associated with reduced risks of venous thrombosis (OR=0.1, 95%CI 0.07-0.14; P=3×10^-43^) and ischemic stroke (OR=0.47, 0.36-0.61; P=2×10^-8^).^36^ MR analysis of our study also revealed a causal relationship between *F11* and CES, and 63.8% of the effect is mediated by FXI. Currently, FXI inhibitors have been developed as potential stroke targets, and their role is evaluated by some ongoing trials.^37–39^ *SCARA5* is a scavenger receptor that exports ferritin-bound iron from circulation to parenchymal tissues, including the heart and brain.^40^ Previous MR studies revealed *SCARA5* genetically lower serum iron which further decreases CES risk.^8,41^. Using multi-omics analysis, we confirmed this result, *SCARA5* was associated with a lower risk of AS (OR [95% CI]: 0.92 [0.87, 0.98]).

To our best knowledge, the other four genes including *ENGASE*, *SPATA20*, *SWAP70*, and *SH3BGRL3* have not been extensively studied in stroke research. And we were able to identify a shared causal variant between a pQTL and stroke for the four genes except for *ENGASE*. And we identified *SPATA20* as associated with the SVS subtype. Moreover, mediation analysis of the effects of these three genes on stroke subtypes indicated potential mechanisms. For example, the indirect effect of *SPATA20* on the risk of SVS via T2D contributes to one-fourth of the total effect (25.2%). and the proportion of the mediation effect of *SWAP70* and *SH3BGRL3* on AIS and AS via SBP and AF is one-tenth, respectively (*SWAP70*: 14.1%; *SH3BGRL3*: 12.2%). However, the mechanisms of the effects of these proteins are not clear yet. *ENGASE* is an enzyme involved in the processing of free oligosaccharides in the cytosol and is associated with glycosylation. Studies have shown protein glycosylation may affect the occurrence of AIS by regulating the progression of atherosclerosis and AF.^42^ *SWAP70* is a guanine nucleotide exchange factor that participates in the regulation of many cellular processes, its role in many diseases is not clear yet, however, studies showed that *SWAP70* is a protective molecule that can suppress the progression of nonalcoholic fatty liver disease by inhibiting hepatic steatosis and inflammation,^43^ and Pathological Cardiac Hypertrophy, et al.^44^ Both of which were associated with stroke occurrence. *SWAP70* also organizes the actin cytoskeleton, which is crucial for phagocytosis.^45^ The role of *SWAP70* in immune function may be related to its protective effect against AIS, and need further study in the future. *SH3BGRL3* may play a role in maintaining cerebrovascular integrity and maintaining the function of cerebral vascular endothelial cells by activating STAT3 signaling, thereby preventing AS and AIS.^46,47^ Little is known about SPATA 20, it is predicted to be located in the extracellular region, and involved in the carbohydrate metabolic process and cell differentiation. Whatever, further studies are needed to confirm the role of these genes and proteins in stroke.

The strengths of our study include the utilization of the largest and most comprehensive human proteome and summary statistics from the most recent GWAS and protein quantitative trait locus (pQTL) datasets to date, and integrative analysis of multi-omics data including GWAS, PWAS, and TWAS. The stroke GWAS summary datasets we used included data for 1,308,064 European descent individuals, nearly three times the size of previous studies (MRGASTROKE n=446, 696). ^7,8^ The plasma proteomic data we used included a total of 7, 213 European Americans, and the number of significant SOMAmers in this study is almost three times that of plasma pQTL studies conducted in the past in the European ancestry sample.^48^ We further identified risk genes through cell enrichment and functional enrichment analysis using single-cell transcriptomes and GO databases. Afterward, we used independent MR analysis to verify PWAS-significant genes. Then we used a colocalization to integrate GWAS data and plasma pQTL using a Bayesian colocalization analysis to explore whether two associated signals are consistent with shared causal variant(s). Finally, we included five common risk factors of stroke in two-step MR analyses to explore the clinical mechanisms of risk genes and explored the significant genes driving GWAS signals at the transcriptional level by leveraging gene expression data.

Our study also has some limitations. First, although the plasma proteomic data we used included SOMAmers for ∼ 5,000 proteins or protein complexes, it does not provide coverage for the entire plasma proteome.^9^ Also, the data did not explore the effects of uncommon and rare variants, as well as complex trans-associations, analysis with substantial discovery on an even larger sample size is likely to need. Second, it is insufficient to elucidate the numerous stroke PWAS-identified loci from genetic and transcriptional levels. Functional genomic approaches are needed to identify the complex molecular mechanisms of stroke. Third, our study mainly focused on European subjects, and caution should be taken when generalizing our results to other ethnicities.

## CONCLUSION

In conclusion, we found causal evidence supporting three classic genes (*MMP12*, *F11*, and *SCARA5*) and three novel genes (SH3BGRL, *SWAP70*, and *SPATA20*) associated with ischemic stroke and their subtypes. Our findings provide genetic evidence underlying ischemic stroke and subtypes, allowing novel therapeutic targets to be further identified.

## Data Availability

All data in the manuscript is truly usable

## Supplemental Material

Tables S1-S10

